# Gamification Enhances User Engagement and Task Performance in Prosthetic Vision Testing

**DOI:** 10.64898/2025.12.20.25342740

**Authors:** Lily M. Turkstra, Byron A. Johnson, Arathy Kartha, Gislin Dagnelie, Michael Beyeler

## Abstract

**Purpose:** Visual function testing in retinal prosthesis users relies on repetitive psychophysical tasks that are cognitively demanding and fatiguing. Gamification may increase engagement, but its effects on perceptual performance in implanted users remain unclear.

**Methods:** Three Argus II users completed circle localization and motion direction discrimination in clinical and gamified versions. Visual stimuli, trial structure, and response requirements were matched within each participant; gamified versions added scoring, background music, and affectively framed end-of-trial auditory feedback. Difficulty and response format were calibrated to individual abilities (8AFC for two participants; 4AFC restricted to cardinal directions for one participant).

**Results:** Gamification improved accuracy and reduced angular error in localization but did not improve motion discrimination. Effects were task-dependent and varied across participants, with reduced precision in the gamified motion task for one user. Participants preferred gamified localization and reported higher enjoyment and sustained attention; responses to gamified motion were mixed.

**Conclusions:** Gamification can influence measured performance and user experience in prosthetic vision testing, but benefits are not universal and depend on task demands and cognitive load, indicating that engagement can affect outcomes in tests often treated as objective.

**Translational relevance:** Personalized, engagement-aware gamified tools with adaptive difficulty may improve the usability and scalability of prosthetic vision assessment and rehabilitation, including at-home training.

## Introduction

Visual neuroprostheses offer a promising approach to restoring functional vision for individuals with profound blindness ^1,2^. Outcomes, however, have varied substantially across device generations. Early retinal systems such as Alpha-IMS and Argus II enabled some users to perform above chance on structured laboratory tasks and to demonstrate improvements on select functional vision measures (e.g., obstacle-related and object-sorting tasks) when the system was on compared with off ^3–10^, but real-world benefit and long-term adoption were highly heterogeneous ^11–13^. In contrast, more recent retinal systems, including the subretinal photovoltaic implant PRIMA and emerging suprachoroidal approaches, have reported substantially improved spatial resolution and task performance in clinical trials ^14,15^, indicating that advances in device design can improve spatial resolution and performance on specific measured tasks in clinical trials.

At the same time, successful outcomes depend not only on technological performance and biotolerability, but also on user engagement, motivation, expectation management, and sustained use. Qualitative work consistently highlights usability burden, training demands, fatigue, and mismatched expectations as major barriers to sustained real-world use, even when device function can be demonstrated in controlled tasks ^12,13,16–18^. These considerations underscore a persistent gap in prosthesis development: ensuring that training, habilitation, and rehabilitation protocols are engaging, accessible, and effective.

Most clinical training and assessment for prosthetic vision relies on standardized psychophysical tasks ^6,19^, including object localization and motion discrimination ^3,7^, which are used to quantify device function and guide parameter adjustments. However, these tasks can be lengthy and cognitively demanding, placing a burden on participants and limiting engagement. In practice, rehabilitation protocols also vary across sites and devices and often lack structured at-home training tools, making it difficult for users to sustain consistent practice or track progress over time ^19,20^. As a result, clinical tests that are intended to provide objective measures of visual function may also be influenced by attention, motivation, and fatigue, complicating interpretation and reducing ecological validity ^21–23^.

One approach to addressing these challenges is to deliberately increase user motivation and engagement during training and assessment. Gamification, defined as the integration of game elements into non-game contexts ^24^, and related “serious game” approaches ^25^ can enhance perceptual learning and visuomotor coordination ^26^. Game-based paradigms have been used effectively in health care settings ^26–28^, specifically in rehabilitation and accessibility ^29–33^. Proposed mechanisms include recruitment of reward and attention systems that may facilitate learning and retention ^34–36^. At the same time, game mechanics can introduce additional cognitive load or distraction, and user preferences vary, suggesting that effectiveness may depend on task demands and individual differences ^37–39^.

Despite this potential, gamification has not been systematically evaluated in the context of retinal prosthesis testing with implanted users. Related approaches have been successfully applied in adjacent domains, including training and assessment for cochlear implant users ^40^, and in studies of simulated prosthetic vision with sighted participants ^41,42^. However, whether gamification alters perceptual performance or user experience in individuals implanted with a retinal prosthesis remains unknown. Here, we asked whether gamifying two standard clinical psychophysical tasks, object localization and motion direction discrimination, influences perceptual performance and engagement in retinal implant users. We quantified performance using accuracy and response precision, and assessed user experience using brief surveys and open-ended probes across standard and gamified task versions. We further explored individual differences in responsiveness to gamification and examined whether adaptive difficulty could help sustain engagement across repeated testing. Understanding these factors is essential for designing personalized training protocols that maximize both usability and rehabilitation outcomes in visual prosthesis users.

## Methods

### Study Participants

This study involved three completely blind participants (one female and two male) with severe retinitis pigmentosa (RP), ranging from 31-90 years at the time of testing (summarized in **Table 1**). Each participant had been chronically implanted with the Argus II retinal prosthesis system prior to enrollment in the present study, following its commercial release and completion of the interventional feasibility trial (clinicaltrials.gov: NCT00407602). The present work involved behavioral task assessments only and did not include surgical intervention or changes to clinical care.

**Table 1.** Demographic and experiential characteristics of our study participants. Because Argus II users are rare and identifiable, ages and implantation durations are reported in ranges to protect privacy.

| Participant | Age | Gender | Years implanted | Device usage | Prior psychophysics experience | Prior gaming experience |
| --- | --- | --- | --- | --- | --- | --- |
| 1 | 51-70 | female | 6-10 | infrequent; primarily during research studies | extensive; frequent studies at Wilmer Eye Institute | none |
| 2 | 51-70 | male | 6-10 | rarely; occasional navigation | extensive; frequent studies at Wilmer Eye Institute | infrequent |
| 3 | 31-50 | male | 1-5 | regularly; at-home use for perceptual tasks | limited; only during follow-ups with Second Sight | extensive |

All psychophysical experiments were conducted in the Ultra Low Vision Laboratory (ULV Lab) at the Lions Vision Research and Rehabilitation Center of the Johns Hopkins Medical Institutions in Baltimore, MD, USA. The study was approved by a centralized Institutional Review Board (WCG IRB) with local institutional oversight at Johns Hopkins University, and was conducted in accordance with the tenets of the Declaration of Helsinki. Participants provided informed consent after receiving an explanation of the study’s nature and potential risks. The consent process was made accessible by reading aloud the form’s text, providing a signature guide to participants, and sharing an electronic version ahead of time ^43^. Medical and research staff were available to answer any participant questions. Participants were compensated at a rate of $20 per hour, including travel time, and were allowed unlimited breaks with the option to discontinue at any time.

Participants’ experience with the Argus II varied. Participants 1 and 2 had prior involvement in psychophysics studies at the ULV Lab. Participant 1 initially used the device for navigation but now primarily engages with it when participating in research studies. Participant 2 previously relied on the implant for navigation and work but has since reduced usage, citing limited benefits and functionality without upgrades. In contrast, Participant 3, despite minimal formal training, reported frequent at-home self-guided perceptual practice with the device.

### Retinal Implant

The Argus II system comprises a 6×10 array of platinum disc electrodes, each 225 *μ*m in diameter, spaced 575 *μ*m apart, and subtending approximately 12×20 degrees of visual angle ^44,45^ (**Figure 1**). The external head-mounted system includes a camera and wireless transmitter mounted on a pair of glasses. The camera captures video input, which a video processing unit (VPU) converts into electrical stimulation patterns for the retinal implant using predefined image processing algorithms.

**Figure 1.**
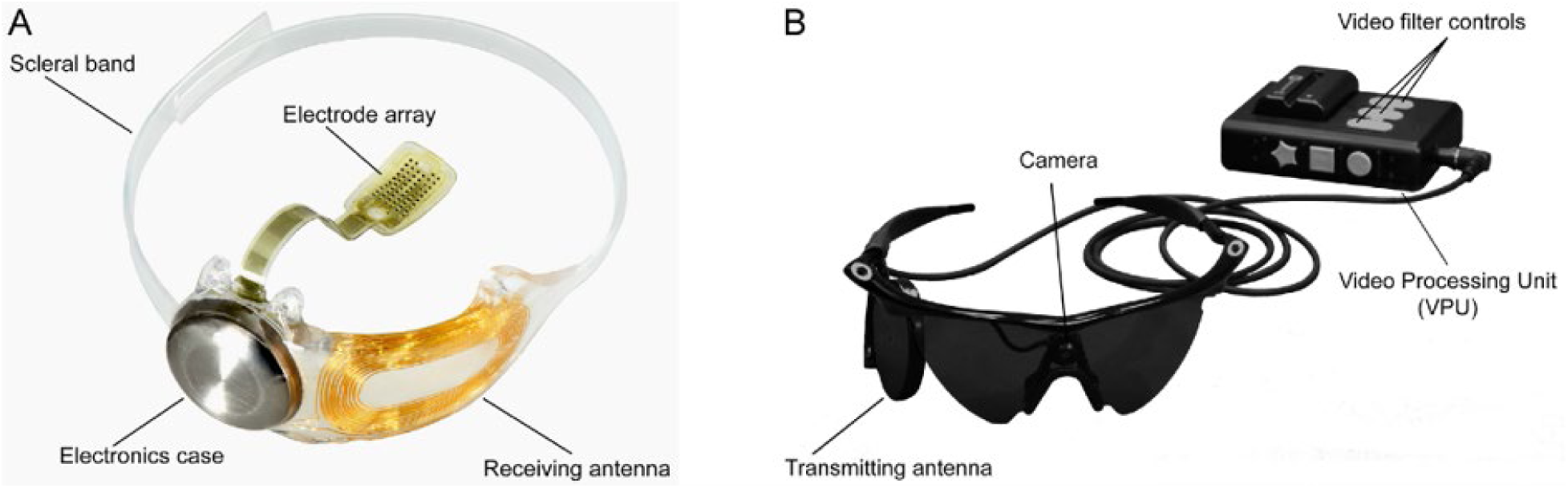
Overview of the Argus II system (reused under CC-BY-ND from ^46^). A) Implanted components include a hermetically sealed electronics enclosure, a receiving antenna secured to the eye with a scleral band and sutures, and a 60-electrode array tacked over the macula. B) External components consist of a glasses-mounted camera connected via cable to a belt- or shoulder-worn video processing unit (VPU).

### Tasks & Gamification

Two psychophysical tasks were selected for gamification: circle localization ^4,7^ and motion direction discrimination ^3,47^ (**Figure 2**), both widely used to diagnostically assess spatial perception and motion processing in Argus II users. While prior studies have demonstrated that prosthesis users perform above chance in these tasks, their accuracy remains highly variable across users and significantly lower than that of sighted individuals performing equivalent tasks under simulated prosthetic vision ^5,48^. Previous work employing these tasks with Argus II and other retinal implant users reports a marked variability in performance between the two, with motion discrimination consistently more difficult than object localization ^48^. These tasks, which are commonly used, differ in their reported difficulties while eliciting performance differences above chance, are therefore well-suited for evaluating the potential benefits of gamification in prosthesis users.

**Figure 2.**
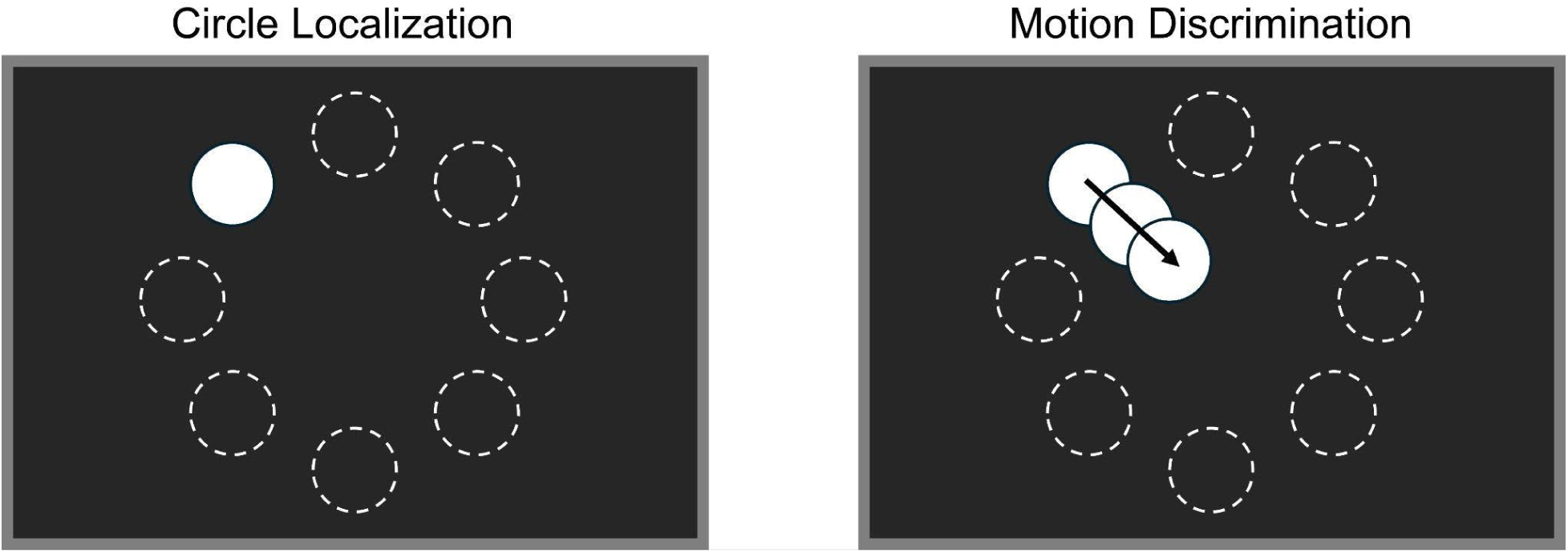
Schematic of the two experimental tasks. (Left) In the circle localization task, the target appeared in one of eight possible locations, and participants indicated its position using a game controller. (Right) In the motion discrimination task, the target’s drift direction was dictated by its starting position, always traveling in a fixed direction across the screen (e.g., a target appearing in the top-left position would always move diagonally down-right). Visual stimuli were identical between clinical and gamified conditions; the gamified condition added auditory and scoring elements (e.g., themed sounds/music and point feedback), while stimulus presentation and response requirements were otherwise unchanged. Dashed circles indicate possible target locations for illustration only; they were not visible during the experiment. Target and screen sizes are not to scale (see Table 3 for actual sizes).

**Table 2.** Examples of auditory feedback phrases used in the gamified condition, categorized by response accuracy. Hits, near misses (± 45°), and misses (>|45°|) triggered different verbal responses, sampled randomly from a database of approximately 25 phrases per category. Phrases were sampled with equal probability throughout the session and delivered only after the response on each trial. Feedback timing and categorical information (hit, near miss, miss) were held constant across clinical and gamified conditions; only the phrasing differed.

| Hit | Near Miss | Miss |
| --- | --- | --- |
| 10 points! | Almost! | Not quite. |
| Ding ding ding, 10 points! | Nearly there! | Keep trying. |
| That was perfect! | Close, but no cigar! | Let's try that again. |
| You're getting good at this! | Just missed it! | No points this time. |
| You're unstoppable! | That was close! | Maybe next time. |
| Easy, isn't it? | Missed it by a hair! | Swing and a miss. |
| Did you secretly practice this? | You almost had it! | Well, at least you're learning. |
| Who knew psychophysics could be fun? | That was really close! | Nope, but you're still a winner to me. |

**Table 3.**
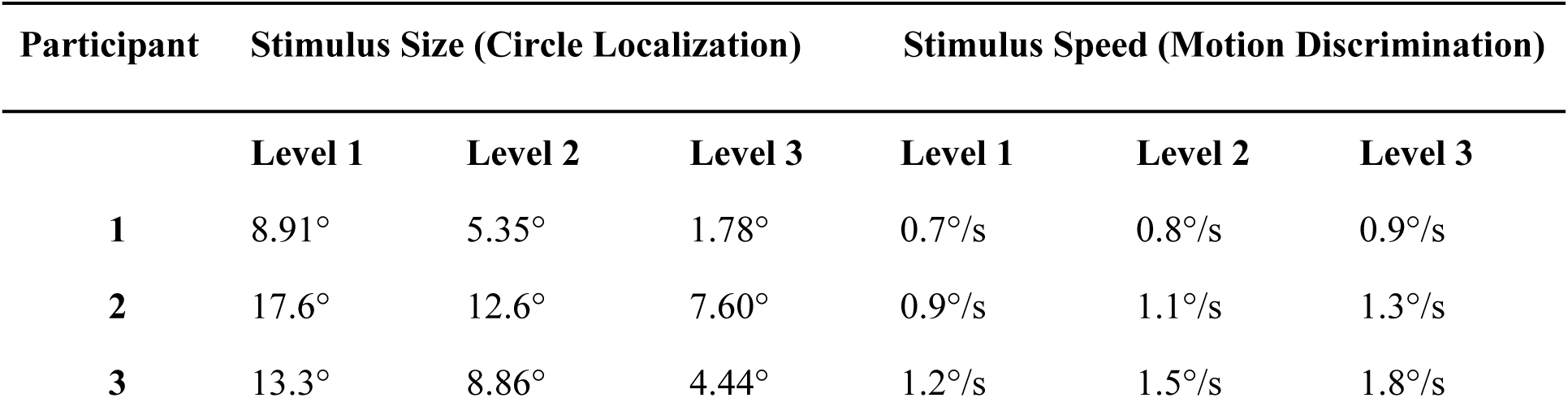
Participant-specific stimulus parameters across three difficulty levels for circle localization and motion direction discrimination (degrees visual angle, dva). In localization, target size decreased with increasing level. In motion discrimination, stimulus speed increased with level; target size was held constant within each participant across motion levels (set from that participant’s Level 1 localization size) to emphasize motion discrimination rather than localization difficulty. Parameters were individualized to reduce the likelihood that condition effects were driven by floor or ceiling performance or mismatched task demands, rather than by task framing.

Gamification was implemented by embedding task-relevant game mechanics into each paradigm while keeping visual stimuli, response requirements, and trial structure identical to the clinical versions. The circle localization task was reframed as a balloon-popping game, where participants had to correctly identify the balloon’s location to “pop” it by indicating its location with a game controller, accompanied by balloon inflation sounds, a popping effect for correct responses, and deflation for incorrect responses. Similarly, the motion direction task was transformed into a fishing game, where correct responses were rewarded with a reeling-in sound to indicate catching a fish, while incorrect responses played a bubbling sound to simulate the fish escaping.

Additional game elements were introduced to enhance engagement, including point-based incentives, background music, and affectively framed auditory feedback. Importantly, feedback timing and informational content were held constant across conditions: in both the standard and gamified tasks, participants received auditory feedback only at the end of each trial, indicating whether the response was correct, near-correct, or incorrect. No continuous or real-time performance feedback was provided during stimulus presentation or response execution.

In the gamified condition, this end-of-trial feedback was delivered using a larger and more varied set of verbal responses (**Table 2**) designed to convey encouragement and engagement (e.g., “You’re getting good at this!”), whereas the standard condition used a small, neutral set of task-oriented feedback phrases (e.g., “Correct” or “Incorrect”). Participants could request their cumulative score at any point during a trial block or session, but scores were not displayed automatically. Scores were computed from response accuracy and angular error. Background music was present only in the gamified condition and differed by task, with a carnival theme for localization and orchestral music for motion discrimination. The goal of these modifications was not to isolate the effects of individual game mechanics, but to assess whether a realistic combination of commonly used gamification elements could increase engagement and motivation when applied to standard visual function testing.

### Experimental Setup

All testing was conducted in a darkened room in the ULV Lab to minimize ambient light interference and optimize contrast for prosthesis users. Participants were seated in front of a 69.6 cm × 39 cm television monitor (Samsung LN-S3251D) at a viewing distance ranging from 42 cm to 59.7 cm, depending on individual comfort and ability to perceive stimuli (Participant 1: 42 cm, Participant 2: 48 cm, Participant 3: 59.7 cm). Stimuli were generated using SimpleXR ^49^, an open-source Unity-based toolbox for virtual and augmented reality (https://github.com/simpleomnia/sxr). The monitor displayed all stimuli, and responses were recorded using a conventional video game controller. Participants (all three right-handed) used their left thumb to indicate stimulus location or direction via the controller’s directional pad and confirmed their responses by pressing a separate button with their right index finger.

To provide a consistent frame of reference, tactile markers (clear tape) were placed at the center and along the edges of the monitor, and did not interfere with stimulus perception. At the beginning of each block, participants were instructed to reorient themselves by feeling the tactile markers. While participants were free to move their heads to scan for stimuli, they were encouraged to avoid excessive touching of the screen to prevent reliance on tactile cues. The monitor height was adjusted for Participant 3 to accommodate their preferred interaction style.

All experiments were conducted with the Argus II device turned on and in standard video mode. The video processing unit (VPU) applied the system’s default contrast enhancement settings without additional image processing.

### Block Structure & Difficulty Progression

To systematically vary difficulty, the circle localization task used progressively smaller targets across levels, while the motion direction task increased stimulus speed. Since Argus II users have highly variable perceptual abilities, absolute stimulus parameters differed across participants. The easiest level for each condition was calibrated individually so that participants could achieve approximately 80% accuracy, ensuring that difficulty progression was meaningful and engaging, while keeping overall task demands comparable.

In the circle localization task, difficulty escalated by progressively shrinking the target; for motion direction discrimination, difficulty increased with stimulus speed. Raw values for each participant’s stimulus parameters are provided in **Table 3**.

In the gamified condition, these difficulty changes were framed as advancing through game levels, reinforcing a sense of progression. Verbal feedback emphasized challenge and achievement, transforming what would otherwise be passive difficulty increments into a more immersive and engaging experience.

### Training & Testing Procedure

The full procedure timeline is outlined in **Figure 3**. Before testing, participants completed a structured training phase to ensure they understood the task and could reliably use the game controller for responses. After familiarizing participants with the desktop setup, training began with a simplified version of each task in which only two response options were available (e.g., left vs. right for localization, upward vs. downward motion for discrimination). This allowed participants to acclimate to the task structure and response mapping before progressing to the full task.

**Figure 3.**
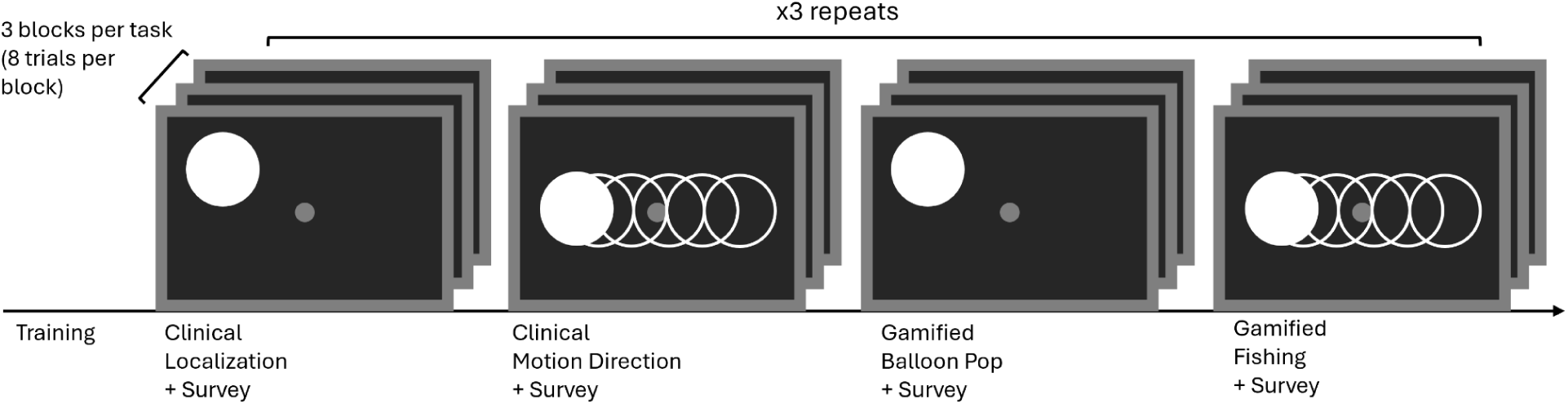
Experimental procedure. Participants first completed a training phase to familiarize themselves with the tasks and response system. Testing alternated between clinical and gamified conditions, with localization and motion discrimination tasks presented in separate blocks. Each task comprised three difficulty levels per condition, with eight trials per block. The full experiment was repeated three times across two sessions (288 trials per participant). The response format was calibrated to individual perceptual abilities (8AFC for Participants 1–2; 4AFC, cardinal directions only, for Participant 3), while trial counts and stimulus presentations were matched between clinical and gamified conditions. Surveys were administered after each condition to assess engagement and experience.

Initially, responses were provided verbally, and once participants demonstrated consistent understanding of the task requirements, they transitioned to using the game controller for all responses. Training continued until both the participant and the experimenter were confident that the task was understood and that responses were reliable.

Following successful completion of the simplified training, participants advanced to the full experimental version of each task. To accommodate individual differences in perceptual abilities, response format was calibrated on a per-participant basis. Participants 1 and 2 completed an eight-alternative forced-choice (8AFC) version of the task, whereas Participant 3 completed a four-alternative forced-choice (4AFC) version restricted to the cardinal directions. In the 4AFC condition, only the four corresponding target locations or motion directions were presented, and only those labels were available as valid responses; oblique directions were neither displayed nor analyzed.

Importantly, the number of trials, stimulus presentations, block structure, and difficulty progression were held constant across participants and were matched between the clinical and gamified conditions, ensuring that differences in performance reflected task framing rather than stimulus exposure or trial count.

### Statistical Analyses

All analyses were performed in R (v4.5.2). Given the small sample size (*N* = 3) and repeated-measures structure, we modeled outcomes at the trial level using mixed-effects models and emphasized effect sizes and uncertainty intervals.

Trial-level accuracy was analyzed using a Bayesian generalized linear mixed model (Bernoulli likelihood; logit link; `brms`). Fixed effects included Condition (Clinical vs. Gamified), Task (Localization vs. Motion), their interaction, and Session, with a random intercept for Participant. To account for the differing number of alternatives across participants (4AFC vs. 8AFC), we included an offset term equal to *logit(1/m)*, where *m* denotes the number of alternatives on each trial. This adjusts the baseline intercept such that a linear predictor of zero corresponds to chance-level performance (1/*m*), allowing the model estimates to represent deviations from chance in log-odds units. We report posterior estimates with 95% credible intervals and considered effects credible when the 95% credible interval excluded zero. Angular error magnitude (absolute signed error, in degrees) was analyzed using a generalized linear mixed model with a Gamma distribution and log link (`glmmTMB`), which is appropriate for non-negative, right-skewed outcomes. The fixed- and random-effects structure matched the accuracy model. To verify that group-level effects were not driven by any single participant and to provide distribution-free within-subject confirmation, we additionally performed participant-level permutation tests (10,000 label permutations) comparing Gamified versus Clinical performance within each task.

## Results

### Individual Differences in Gamification Effects

Given the substantial variability in prosthetic vision outcomes, we first examined the impact of gamification at the individual level using non-parametric permutation tests (10,000 resamples). The effectiveness of gamification varied considerably across participants, revealing distinct response profiles in both task accuracy (**Figure 4A-C**) and perceptual precision (**Figure 4D-F**).

**Figure 4.**
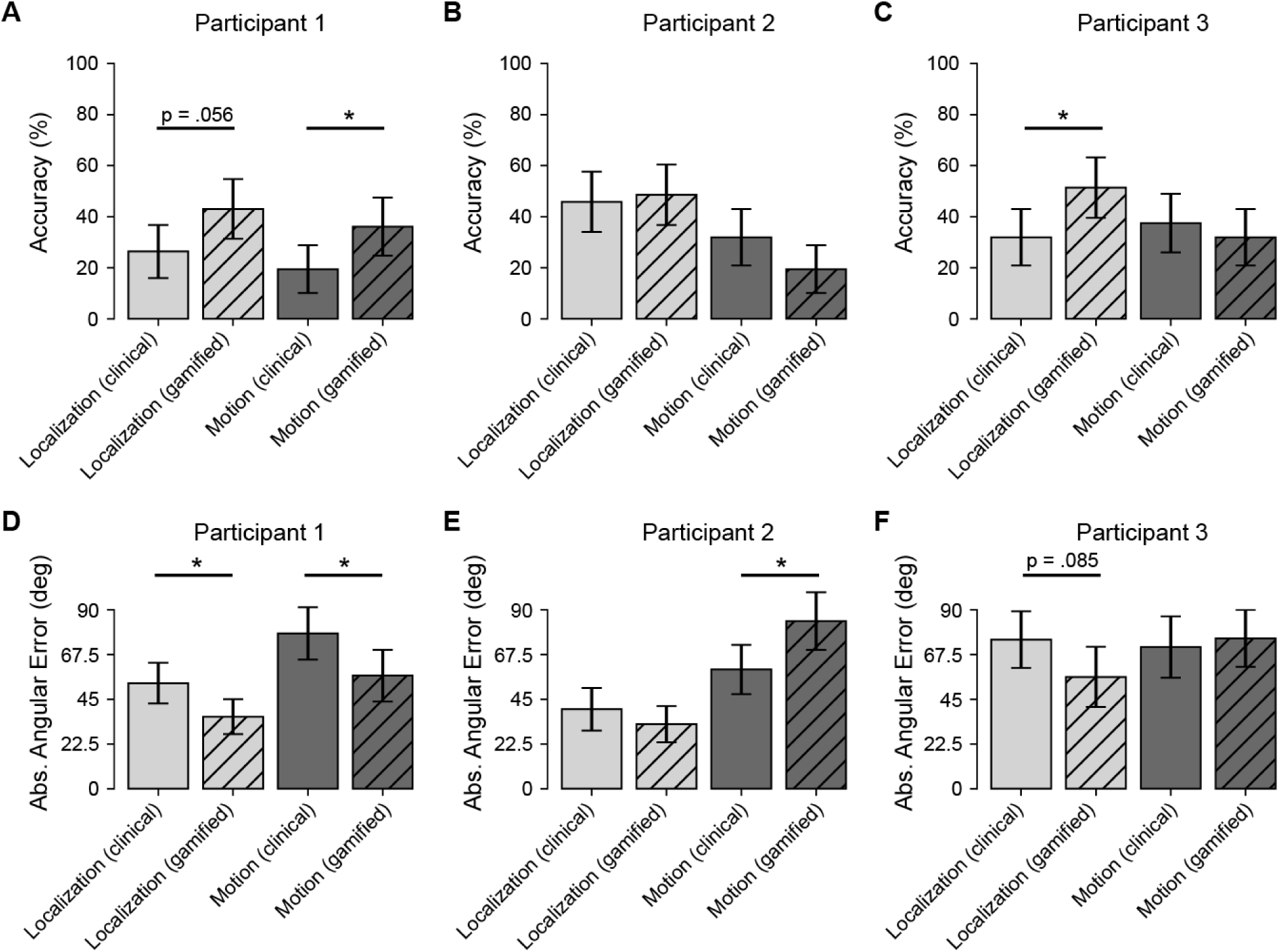
Individual participant performance across clinical and gamified conditions. Panels A–C show trial accuracy (%) for Participants 1–3, and panels D–F show absolute angular error (degrees) for the same participants. Each panel compares the four within-subject conditions: localization (clinical), localization (gamified), motion discrimination (clinical), and motion discrimination (gamified). Error bars indicate 95% confidence intervals. Asterisks denote participant-level permutation test results for the gamified vs. clinical contrast within each task (*p* < .05). Overall, gamification was associated with higher localization accuracy and lower localization error in most participants, whereas effects on motion discrimination were smaller and more heterogeneous across individuals.

Participant 1 demonstrated the broadest benefit from the gamified interface, showing consistent improvements across both metrics. In terms of accuracy, they showed a significant improvement in the motion discrimination task (*p* = .041) and a marginally significant improvement in the localization task (*p* = .056). Crucially, this user also exhibited significant gains in perceptual precision. Permutation tests on mean absolute angular error revealed that gamification significantly reduced error magnitudes in both localization (*p* = .016) and motion discrimination (*p* = .028), reducing mean error by approximately 17° and 21°, respectively. This suggests that for high-performing users, gamification can enhance both the reliability of target selection and the spatial precision of their responses.

Participant 3 exhibited a task-specific benefit limited to static localization. Gamification drove a significant improvement in localization accuracy (*p* = .029) and a trend toward improved precision (p = .085), with a mean error reduction of 19°. However, these benefits did not extend to the dynamic motion task, where both accuracy (*p* = .59) and precision (*p* = .72) remained comparable to clinical baseline performance.

In contrast, Participant 2 illustrated the potential cognitive costs of gamification. While their accuracy remained stable across conditions (*p* > .1), their precision in the motion task significantly worsened in the gamified condition (*p* = .014), with mean error increasing by approximately 24° compared to the clinical baseline. This significant decline suggests that for some users, the additional auditory and cognitive load of the gamified motion task (i.e., the “fishing” metaphor) may have acted as a distracter rather than a motivator, interfering with the processing of the visual motion signal.

### Gamification Improves Accuracy in the Localization Task

To assess whether these individual benefits translated to a consistent group-level effect, we employed a Bayesian Generalized Linear Mixed Model (GLMM). Consistent with the individual findings, the model revealed a significant main effect of gamification on accuracy in the localization task (*p* = 0.51, 95% CI [0.14, 0.89]). Post-hoc contrasts indicated that gamification increased the odds of a correct response by approximately 1.68 times compared to the clinical condition.

However, this benefit was task-dependent at the group level. A significant Condition x Task interaction (*p* = -0.52, 95% CI [-1.06, 0.02]) indicated that the accuracy gains observed in localization did not consistently extend to the motion discrimination task. For motion discrimination, group-level performance remained comparable to the clinical baseline (OR ≈ 1.01, 95% CI [0.64, 1.46]). This suggests that while gamification reliably boosts performance in spatial localization, its ability to overcome the sensory challenges of motion processing is highly user-dependent, as evidenced by the contrasting outcomes of Participant 1 and Participant 2.

### Precision and Error Magnitude

We further analyzed perceptual precision at the group level using a Gamma GLMM on absolute angular error. Mirroring the accuracy results, we observed a trend toward improved precision in the gamified localization task (*p* = .077). On average, angular errors in the clinical condition were 1.34 times larger than in the gamified condition. No significant difference in precision was observed for the motion task at the group level (*p* = .83), reflecting the high inter-subject variability where one participant improved significantly while another worsened.

### Absence of Learning Effects

To ensure that the observed improvements were due to the gamified interface rather than task familiarity or practice, both the accuracy (Bayesian) and precision (Gamma) models controlled for session effects. No significant learning effects were observed for accuracy (Session 2: *p* = -0.16; Session 3: *p* ≈ 0.00) or precision (Session 2: p = 0.51; Session 3: *p* = 0.89). This confirms that performance changes (both positive and negative) were driven by the task structure itself rather than sequential learning over the course of the experiment.

### Effect of Gamification on Enjoyment and Perceived Difficulty

To complement the performance analysis, participants rated their enjoyment (1-10) and ease of use, with higher values indicating greater enjoyment and greater ease (**Table 4**). Additional survey items assessed sustained attention and motivation (Yes/No/Neutral), as well as overall task preference.

**Table 4:** Changes in enjoyment (↑higher is better) and perceived difficulty (↓lower is better) between gamified and clinical conditions. Ratings were originally provided on a 1–10 scale, with higher values indicating greater enjoyment or greater difficulty. Reported values reflect the difference (Gamified – Clinical) for each participant and task, where positive values indicate increased enjoyment and difficulty in the gamified condition.

| Participant | Task | $\Delta$ Enjoyment ↑<br>(Gamified - Clinical) | $\Delta$ Difficulty ↓<br>(Gamified - Clinical) |
| --- | --- | --- | --- |
| 1 | Localization | +2 | -2 |
| 1 | Motion | +5 | -4 |
| 2 | Localization | +1 | 0 |
| 2 | Motion | -2 | +1 |
| 3 | Localization | +4 | -2 |
| 3 | Motion | -1 | 0 |

On average, gamified tasks were rated 1.5 points higher in enjoyment and 1.17 points lower in difficulty compared to their clinical counterparts, indicating that gamification not only increased engagement but also made tasks feel easier. All participants preferred the gamified localization task, while opinions on the motion task were mixed, suggesting that the impact of gamification may depend on task type. As Participant 3 remarked:

> *“I was having fun because I was doing better - or I was doing better because I was having fun?”*

Survey responses provided insight into engagement and motivation across conditions. All participants reported being motivated in both the clinical and gamified tasks, except for Participant 1, who rated motivation as neutral in clinical localization. Similarly, all participants indicated that the tasks sustained their attention, except for Participant 1, who reported that clinical localization failed to do so. While gamification did not universally increase motivation, it helped maintain engagement, particularly in tasks that might otherwise feel monotonous.

Reactions to the motion task were mixed. When asked whether gamification would encourage practice, Participant 2 responded:

> *“Yeah, probably … not so much the fishing game, the other game was fine. The fishing game is kind of like teasing you… ‘Better luck next time’ or ‘You missed it.’ One way or the other, a beep or bang or a pop makes it a little more interesting, like a pinball machine.”*

Notably, both tasks used the same auditory feedback system, suggesting differences in perception rather than content.

Competitiveness also shaped participant experiences. Participant 3 expressed frustration:

> *“You would think I would know how to do this by now. Come on, I’ve had the chip for five years!”*

Yet, he also saw the challenge as motivating:

> *“I’ve always been competitive, so the better I do, the happier I am playing it. It frustrates me when I don’t get the high score I should. It can drive you nuts to not catch the fish, but at the same time, just by doing the right head movements, I do find benefit from catching fish and popping balloons.”*

In sum, gamification provided both encouragement and, at times, frustration, especially when expectations of improvement were unmet.

## Discussion

Vision rehabilitation for prosthesis users remains a significant challenge, and standard clinical assessments may not fully capture real-world functional abilities. Here, we tested whether gamification can influence perceptual performance and user experience during prosthetic vision testing. Gamification improved accuracy and reduced angular error in the localization task, consistent with the possibility that engagement influenced measured precision. In contrast, gamification did not improve, and may have hindered, motion direction discrimination, where errors remained higher. Together, these results indicate that gamification can enhance performance in some contexts but is not universally beneficial. No learning effects were observed across trials or sessions, supporting the interpretation that performance differences reflected task framing rather than short-term practice.

### Gamification Enhances Performance, But Its Effects Are Task-Dependent

All participants preferred the gamified localization task over its clinical counterpart, reporting higher enjoyment and same or lower perceived difficulty (**Table 4**). However, the gamified motion task remained challenging, consistent with prior findings that motion discrimination is inherently more difficult and requires more training ^50^. This suggests that while gamification can increase engagement, its impact on performance depends on task complexity.

Crucially, gamification did not just improve subjective experience, but enhanced objective performance in localization (**Figures 4–5**). Therefore, traditional acuity and function tests may not fully capture attentional and cognitive factors influencing prosthesis use. Prior research suggests that engaging tasks activate attentional networks and promote neuroplasticity ^34,36^, which may explain the observed improvements in localization. However, the absence of gains in the motion task suggests that gamification alone may not overcome task-intrinsic difficulty and, in some cases, may impose additional cognitive load. This pattern was also evident at the individual level, reinforcing that mechanics that support engagement for one user may distract another.

### Tailoring Gamified Training to Individual Needs

Gamification’s effectiveness varied across participants (**Figure 4**), reinforcing that one-size-fits-all approaches may not be optimal for clinical applications. Individual differences (e.g., prior training, head-scanning techniques, and baseline perceptual abilities) play a critical role in task performance ^3^. Participants 1 and 2 had extensive experience with head scanning, while Participant 3 had little prior training and required additional guidance, which may have influenced their performance. Future gamified tasks should adapt to individual perceptual strategies, ensuring that engagement-enhancing elements do not introduce unnecessary distractions ^33^. In practice, this suggests that future at-home tools should combine personalization with safeguards that prevent gamification from becoming a barrier to task completion.

### Limitations and Future Directions

While this study highlights the potential of gamification for perceptual testing and training, several limitations should be considered. First, the small sample size (*N* = 3) limits generalizability, reflecting the rarity of Argus II users available for research. Although a case-study approach provides valuable insight into individual variability and the need for customization, larger cohorts will be necessary to systematically examine factors such as age, prior gaming experience, and training history. Future work should also include users of other retinal and cortical prostheses to assess whether these findings extend beyond the Argus II system.

Second, the study was not designed to assess longer-term learning or neuroplasticity. Testing occurred across two sessions with a limited training dose, which constrains the ability to detect practice effects or durable changes in performance, fatigue tolerance, or attention. Accordingly, the absence of learning effects should not be interpreted as evidence that learning does not occur in retinal implant users. Longer-term studies incorporating extended training, retention tests, and transfer to more functional tasks will be required to determine whether gamification supports sustained improvements beyond immediate task performance.

Finally, gamification elements were implemented in combination to reflect realistic clinical use, rather than to isolate individual mechanics. As a result, the contribution of specific elements such as scoring, music, or affective feedback cannot be disentangled. While the gamified localization task was consistently preferred, the motion discrimination task received mixed feedback, suggesting that some design choices may introduce distraction or cognitive load in already demanding tasks. Future studies should therefore systematically evaluate individual game mechanics and explicitly measure cognitive load and fatigue alongside performance.

### The Potential for Personalized, At-Home Gamified Training

Despite these limitations, gamified perceptual tasks could provide a scalable option for at-home vision training, particularly for users who lack access to in-clinic rehabilitation. However, training design must carefully manage how users experience success and failure. Prior qualitative work suggests that failures are often internalized by users, whereas successes are frequently attributed to the device or system rather than to the user’s own skill ^12,13^. This asymmetry underscores the importance of adaptive difficulty and feedback design that preserves a high likelihood of success while still providing meaningful challenge. Training paradigms that emphasize frequent, achievable successes may help sustain engagement and reinforce user confidence, while reducing discouragement that can arise from repeated failure in demanding tasks.

A personalized gamified system could support rehabilitation by dynamically adjusting difficulty based on a user’s scanning behavior, skill level, and perceptual strategies, thereby increasing task demands gradually without becoming overwhelming. It could also enable self-paced practice that preserves user agency while remaining clinically grounded through structured benchmarks defined collaboratively with clinicians and users, helping align expectations for progress. Finally, it could increase accessibility by providing consvistent training opportunities for users who cannot regularly attend specialized rehabilitation clinics or who benefit from customized motivational supports to sustain adherence.

Beyond engagement, gamification may serve as a practical vehicle for delivering sufficient training dose, which is a prerequisite for perceptual learning in many visual tasks. More broadly, models of attention and reward suggest that reinforcement mechanisms could be leveraged to support sustained participation and learning ^34–36^, although these longer-term effects remain to be tested directly in implanted users ^22^. Future work should therefore evaluate extended at-home deployment with retention and transfer measures to determine whether improved engagement translates into durable functional gains.

Taken together, our findings suggest that gamification can meaningfully alter both user experience and measured performance in standard prosthetic vision tasks. Incorporating engagement-aware, adaptive task designs into clinical testing and rehabilitation may improve the reliability of visual function assessment and help more users access effective, sustainable training.

## Data Availability

The data that support the findings of this study are available from the corresponding author upon reasonable request. Due to the small number of participants and the risk of re-identification, data are not publicly available.

